# In-depth phenotyping of human peripheral blood mononuclear cells in convalescent COVID-19 patients following a mild versus severe disease course

**DOI:** 10.1101/2020.05.25.20112763

**Authors:** Chang-Feng Chu, Florian Sabath, Shan Sun, Ying-Yin Chao, Christina E. Zielinski

**Author notes:** **Corresponding author:** Prof. Dr. med. Christina Zielinski, Institute of Virology, Center for Translational Cancer Research (TranslaTUM), Technical University of Munich, Germany.

## Abstract

**Background:** Covid-19, the disease caused by infection with SARS-CoV-2, has developed to a pandemic causing more than 239, 000 deaths worldwide as of 6th May according to the World Health Organization (WHO). It presents with a highly variable disease course ranging from a large proportion of asymptomatic cases to severe respiratory failure in 17-29% of cases even in the absence of apparent comorbidities ^1, 2^. This implies a diverse host immune response to SARS-CoV-2. The immunological characteristics underlying these divergent disease courses, however, still remain elusive. While insights into abrogations of innate immunity begin to emerge, adaptive immune responses towards SARS-CoV-2 are poorly investigated, although they serve as immune signatures of protection and vaccine responses. We therefore set out to characterize immune signatures of convalescent COVID-19 patients stratified according to their disease severity.

**Methods:** We performed high-dimensional flow cytometric profiling of peripheral blood mononuclear cells of convalescent COVID-19 patients who we stratified according to their disease severity by a physician-assisted questionnaire based assessment of COVID-19 symptoms.

**Results:** Surprisingly, we did not observe any difference in the relative proportions of any major immune cell type in convalescent patients presenting with different severity of COVID-19 disease except for a reduction in monocytes. The frequency of T_naive_ T cells was significantly reduced in CD4^+^ and CD8^+^ T cells, whereas other T cell differentiations states (T_CM_, T_EM_, T_EMRA_) remained relatively unaffected by COVID-19 severity as assessed approximately two weeks after infection.

**Conclusions:** In our COVID-19 patient cohort, which is characterized by absence of comorbidities and therapeutic interventions other than symptomatic antipyretics, the immunophenotype is similar irrespective of a highly variable disease severity. Convalescence is therefore associated with a rather uniform immune signature. Abrogations, which were previously identified in the innate and adaptive immune compartment of COVID-19 patients should be scrutinized for direct associations with a preconditioned immune system shaped and made vulnerable for SARS-CoV-2 by preexisting comorbidities.

## Introduction

The pandemic caused by SARS-CoV-2 is a global health emergency. The World Health Organization (WHO) declared the outbreak with this novel virus a pandemic. As of now (23^rd^ May 2020), around 5 million people are affected by this virus, with an estimated 3-4 % case-fatality rate with large regional variations reaching up to 10% (WHO status report)^3^. Countries all over the world are currently at unprecedented speed installing regulatory measures to prevent human-to-human viral spread. SARS-CoV-2 causes the disease COVID-19, which is characterized by severe respiratory symptoms. Most patients infected with SARS-CoV-2 present with mild to moderate symptoms such as fever, fatigue, cough and anosmia. A small number of patients (17%), however, progress to severe disease courses characterized by acute respiratory distress syndrome that might ultimately lead to multiple organ failure and death ^1^. Comorbidities, such as cardiovascular disease, diabetes, hypertension, old age and immunosuppressive therapies have been shown to predispose to severe COVID-19 by epidemiological investigations in a large proportion of patients ^1, 4^ But variable severity in the disease course has also been reported even in young and healthy patients ^5–9^. Asymptomatic infected patients pose a great threat to our society since their activity radius increases viral spread. It remains enigmatic what factors predispose to mild versus severe disease in the absence of apparent comorbidities.

The pathogenesis of COVID-19 is strongly associated with a profound impact of host immune responses to the virus. Severe respiratory distress syndrome is associated with a cytokine storm ^10, 11^. Proinflammatory cytokines such as IL-2, TNF-α, GM-CSF and IL-6, IL-1β, IL-8, G-CSF and others have been detected in the plasma of patients with severe illness ^2^ ^12^. Cytokine release is less pronounced in settings of mild disease courses indicating a correlation with clinical features rather than infection with the virus itself ^13^. The innate immune compartment is rapidly activated resulting in cytokine release and tissue damage. This has initiated clinical trials with specific innate cytokine blocking drugs, such as tocilizumab (IL-6R blocking antibody). They demonstrated improvement in respiratory and laboratory parameters^14, 15^. ACE2, the entry receptor of SARS-CoV-2, has been reported to be expressed in several innate immune cell types such as monocytes ^16–18^. Despite the cross-talk of innate and adaptive immunity, little is known about T cell abrogations in COVID-19 until now. In particular, their differential impact on mild versus severe disease remains elusive.

T cells participate in viral clearance through antigen specific cytotoxicity and B cell help ^19, 20^. Their polyfunctional cytokine release contributes to an overall immune activation with downstream inflammatory actions on immune and stromal cells ^21, 22^. An individual’s T cell signature could either predispose or protect from infection or severe COVID-19 illness. The T cell post-activation fingerprint represents an immune signature of protection and is expected to correspond to future vaccination success. Dissecting human T cell responses to SARS-CoV-2 will therefore yield predictive markers for the disease course as well as information on the effect of the viral infection on adaptive immunity. Recently, it has been shown that lymphopenia in both the CD8 and CD4 T cell compartment correlates with severe disease ^13, 23, 24^. T cells displayed increased activation as judged by HLA-DR upregulation ^13, 25^. But also the upregulation of inhibitory receptors such as NKG2A, PD-1 and TIM-3 has been reported, implying T cell exhaustion in response to SARS-Cov2 infection in settings of severe illness ^25^ ^27^ Cytokine expression in T cells from COVID-19 patients showed variable results. The percentage of IFN-γ producing CD4^+^ T cells was increased in severely affected COVID-19 patients, in line with the previously reported contribution of IFN-γ to the cytokine storm in SARS patients ^28^. Others reported IFN-γ to be unaltered or even decreased as part of reduced functional T cell diversity ^26, 29–31^. Other T cell related cytokines showed increased expression levels upon analysis in the serum ^27, 32^. These results imply that productive T cell effector function might precede a state of T cell exhaustion.

In this study we set out to identify immune signatures of mild versus severe COVID-19. We therefore implemented a high-dimensional analytical approach to characterize the composition of peripheral blood mononuclear cells with a focus on T cell differentiation states in convalescent COVID-19 patients stratified according to the severity of their disease course. Importantly, our patient cohort excludes study participants with comorbidities, who might have skewed previous studies towards a more severe disease course and to immune signatures characteristic of the underlying comorbidity and baseline medication.

## Results

### Characterization of the COVID-19 patient cohort

Subjects were recruited after announcement of the study via social media (Facebook site of Klinikum r.d. Isar). More than 7000 shares were recorded by the end of the patient recruitment. 74 patients were randomly selected based on the criteria of a positive SARS-CoV-2-RNA viral test result, lack of comorbidities and balanced sex distribution (37 female, 37 male patients) **(Figure 1a)**. The mean age of our patient cohort was 47 years ranging from 19 to 80 **(Figure 1b)**. Blood donations were performed approximately two weeks after an anamnestic infection event and one to three days after quarantine. At the time of blood donation, no remaining disease symptoms except for occasional fatigue were recorded in any of the patients.

**Figure 1.**
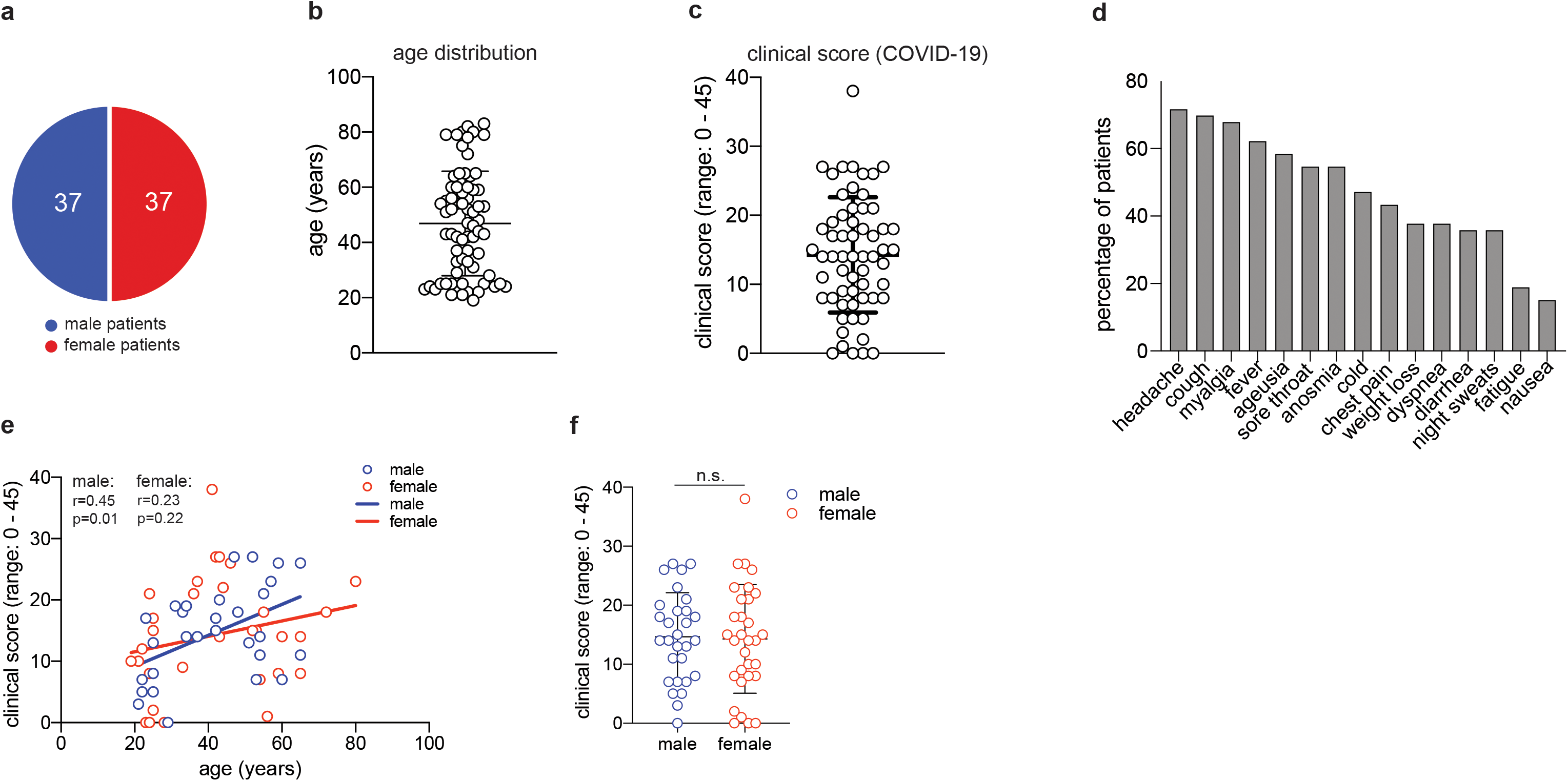
Characterization of COVID-19 patient cohort. **a**, distribution of male and female study participants. **b**, distribution of the age of all study participants (mean ± SEM, 47.3 ± 2.2). **c**, clinical score for each patient (range:0-38) as assessed by a physician assisted questionnaire with each symptom score (range: 0-3) contributing to the final score (mean ± SEM, 14.5 ± 1.1). 60 patient provided clinical metadata. **d**, frequency of patients presenting with the symptoms assessed in the COVID-19 questionnaire. **e**, Spearman correlation r between age and clinical score for male and female patients, respectively. **f**, clinical scores as in c for male and female patients (unpaired student’s t test, n.s. not significant) (mean ± SEM; male: 14.7 ± 1.4; female: mean 14.3 ± 1.7).

We established a clinical COVID-19 score including 15 symptoms this disease has been reported to present with. Quantitative scoring of each symptom (range 0-3) allowed to achieve a final COVID-19 severity score with a mean of 14,5 in our cohort (range: 0-45) **(Figure 1c)**. More than 50% of all patients presented with unspecific symptoms such as headache, cough, myalgia, fever and a sore throat. But also more than 50% of all patients presented with chemosensory alterations such as ageusia and anosmia, which have been proposed to be more characteristic of SARS-CoV-2 infection and to even occur in otherwise asymptomatic infections **(Figure 1d)** ^33^. In our patient cohort the chemosensory loss of taste and smell was consistently associated with other symptoms and always occurred very early in COVID-19 *(not shown)*. Increased age has previously been reported to represent an independent risk factor for severe COVID-19 disease course and death ^6^. Accordingly, we observed slightly higher disease scores in correlation with higher age in men. Interestingly, this age-severity correlation was not significant for women **(Figure 1e)**. Although men have occasionally been reported to develop more severe COVID-19, severity between men and women was equally distributed in our cohort **(Figure 1f)**.

In sum, our patient cohort covered a wide range of COVID-19 severity in the absence of comorbidities and immunosuppression, thus excluding critical predictive markers and immunological abrogations that have previously been correlated with a severe disease course.

### Correlation of immunophenotypes with COVID-19 severity

There is substantial evidence that a dysregulated host immune response plays a critical role in disease severity in several viral infections including SARS-CoV-2. Comorbidities and associated therapies might predispose an individual’s immune system to a more severe disease course and even death as suggested by multiple previous studies ^34^ However, how SARS-CoV-2 infection translates into a mild versus severe disease course in healthy individuals has remained elusive. The host’s response to infection is mirrored by its immune signatures. We therefore set out to characterize the cellular immune cell composition in our convalescent COVID-19 patient cohort and to correlate the immunophenotype with disease severity.

We first quantified the relative proportions of major immune cell types by multi-color flow cytometry of fresh PBMC and correlated them to the respective COVID-19 disease scores of each individual patient **(Figure 2)**. Surprisingly, we did not observe any change in the frequency of CD3^+^ T cells within the lymphocyte population over a wide range of disease severities. This also applied to the frequency of CD4^+^ and CD8^+^ αβ T cells subsets, Treg cells as well as γδ-T cells within the CD3^+^ T cell population. Relative CD19^+^ B cell and CD56^+^ NK cell frequencies within the lymphocyte population were also unaffected by disease severity. The proportion of myeloid cells within the PBMC did, however, decline in correlation with increased COVID-19 (r=0.29, p=0.03). This was also the case for the fraction of CD14^+^ monocytes within the myeloid cell population (r=0.28, p=0.03). We then excluded patients aged 60 and older to test if absence of this independent risk factor, as we showed in the case of men, would explain the negative correlation with myeloid cells or even unravel any new correlation with immune cell frequencies. In fact, the same results could be recapitulated with a patient cohort spanning the age of 19 to 59 (mean 39 years) (data not shown). In sum, this demonstrates absence of any major perturbations in the relative frequencies of the main immune cell types, in particular adaptive immune cells, over a wide range of COVID-19 severity in patients without relevant comorbidities. Only myeloid cells, and in particular CD14^+^ monocytes, correlated negatively with COVID-19 disease severity.

**Figure 2.**
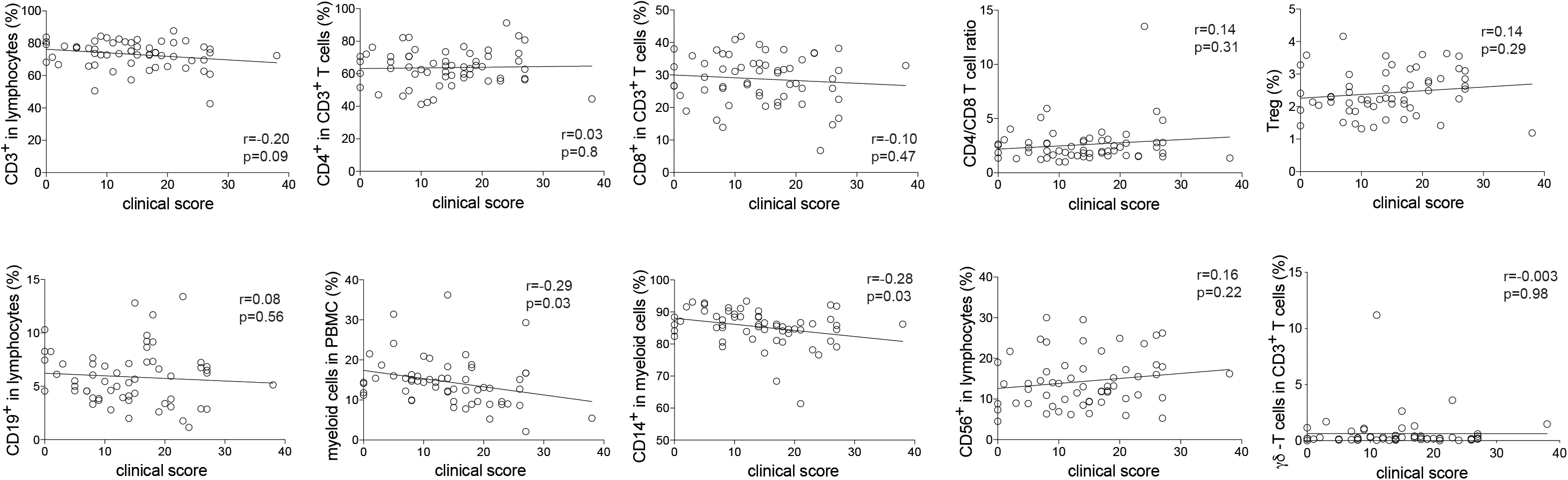
Correlation of immunophenotypes with COVID-19 severity. Flow cytometry of *ex vivo* isolated PBMC gated on the indicated subpopulations. Each circle represents one patient. r, Pearson correlation coefficient, p, p-value

### Correlation of COVID-19 severity with T cell differentiation states

Memory T cells represent the repository of an individual’s antigen experience ^21^. A gradual loss of antigen inexperienced naïve T cells over a lifetime in accordance with a history of antigenic challenges is associated with increased frequencies of memory and effector T cells^35^.The memory T cell pool contains distinct populations of central and effector memory T cells that differ in their homing capacities, functions and history of antigen encounters ^20, 36^. Evidence of T cell activation in both CD4^+^ and CD8^+^ T cells has recently been reported in COVID-19 patients ^13, 37^ We therefore decided to investigate the distribution of distinct T cell differentiation states in the CD4^+^ and CD8^+^ T cell compartment in patients following SARS-CoV-2 infection. Naïve (T_Naive_), central memory (T_CM_), effector memory (T_EM_) and terminally differentiated effector memory cells expressing CD45RA (T_EMRA_) were identified based on the differential expression of CD45RA and CCR7 within CD4^+^ and CD8^+^ CD3^+^ T cells (CD19^-^, CD56^-^, γδ TCR) as described before ^36^. Interestingly, we observed that reduced frequencies of T_Naive_ cells correlated with more severe COVID-19 disease scores in convalescent patients in both the CD4^+^ and CD8^+^ T cell compartments **(Figure 3a,b)**. In CD8^+^ but not CD4^+^ T cells, this was associated with a concomitant increase in T_EM_ cells. T_CM_ and T_EMRA_ cell frequencies did not differ upon correlation with disease scores. Taken together, the distribution of distinct T cell differentiation states in early convalescent COVID-19 patients was overall similar across disease severity scores, with significant changes mainly observable within T_Naive_ cells.

**Figure 3.**
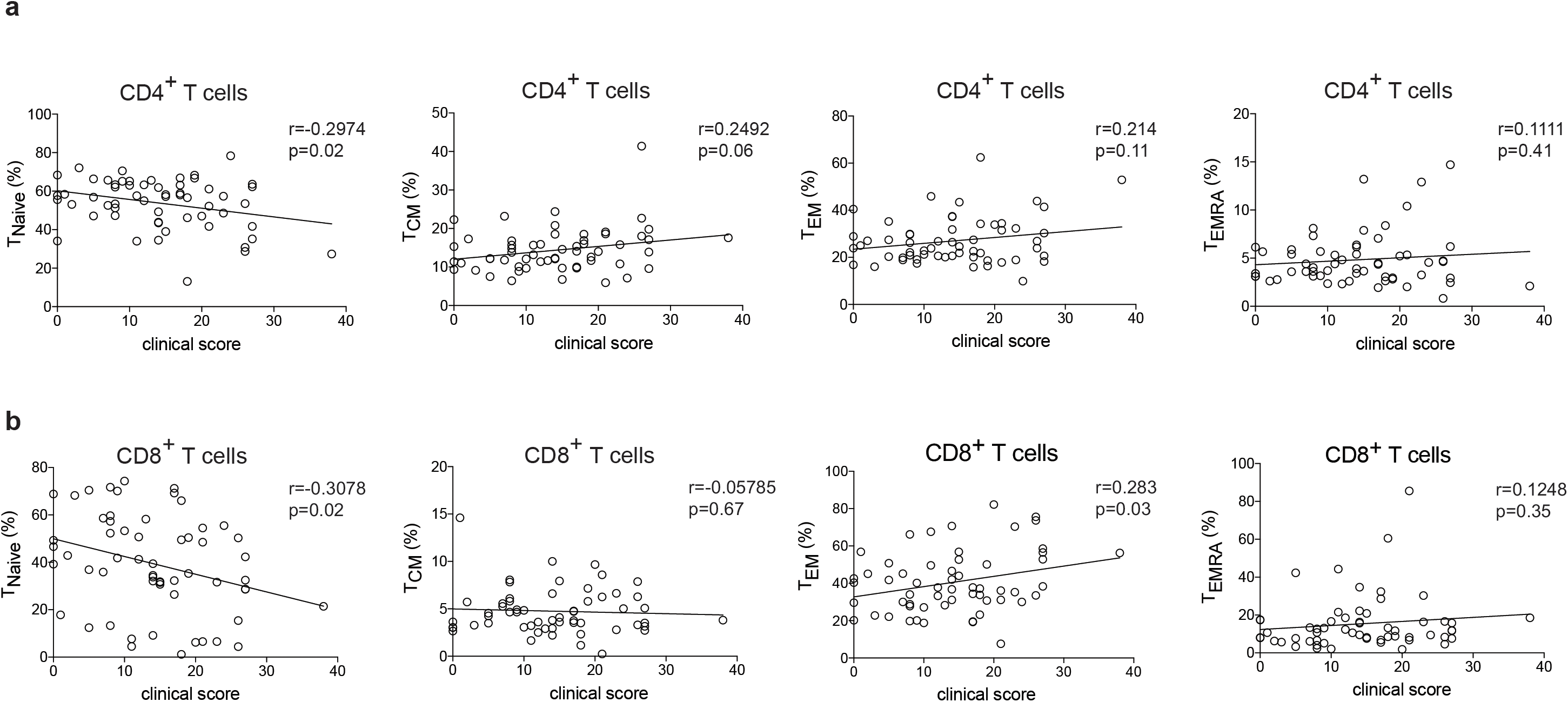
Correlation of COVID-19 severity with T cell differentiation states. Flow cytometry of *ex vivo* isolated PBMC gated on CD3^+^ CD19^-^ CD56^-^ γδ-T cell, CD8^+^/CD4^+^ T cells. T_Naive_: CCR7^+^CD45RA^+^, T_CM_: CCR7^+^CD45RA, T_EM_: CCR7CD45RA^-^, T_EMRA_: CCR7^-^ CD45RA^+^. r, Pearson correlation coefficient, p, p-value

## Discussion

The SARS-CoV-2 pandemic has stirred research efforts with tremendous speed and dedication all over the world. Several studies, which investigated innate and adaptive immunity, have also been initiated ^38, 39^. Comparisons are complicated by variable patient cohorts, which differ in age, sex, time after infection, disease course, comorbidities and medical treatment. We have therefore set out to provide a patient cohort that excludes previously identified risk factors for a severe COVID-19 disease course. Importantly, patients were all investigated at similar timepoints defined by the end of quarantine, which was legally imposed by the German government for two weeks after RT-PCR confirmed diagnosis of SARS-CoV-2 infection. Despite a wide range of disease severity, our patient cohort presented with overall very similar convalescent immunophenotypes that did not correlate with disease scores. The immunophenotypes were reproducible in our COVID-19 cohort after exclusion of patients older than 60 years of age (mean age 39 years) and absence of relevant comorbidities, medication and immunosuppression. Although we did not perform a direct comparison to healthy PBMC samples, the immunophenotypes, which we obtained, were within the physiological ranges reported before, thus excluding any major immune perturbations following SARS-CoV-2 infection. Together, this indicates stability of the immune composition in early convalescent patients over a wide range of disease severities.

Significant alterations were, however, reported within the myeloid cell comportment, which demonstrated reduced cells and, in particular, reduced frequencies of CD14^+^ monocytes in correlation with increased disease severity. This could be due to activation triggered loss of these short lived innate immune cells and would be consistent with the cytokine storm that is mainly composed of innate cytokines such as IL-6 ^39, 40^. Alternatively, monocytes could still be sequestered in peripheral organs, such as the lung, at this early timepoint after clinical recovery in severely affected individuals.

T cells specific for SARS-CoV2 have recently been detected within T_CM_, T_EM_ and T_EMRA_ cells, but their characteristics and role in COVID-19 pathogenesis remained undefined ^41^. Interestingly, we observed reduced frequencies of naïve T cells upon increased COVID-19 severity. This could be due to antigen specific stimulation with SARS-CoV2 antigens and recruitment into the memory T cell pool. This would be in line with the concomitant increase in CD8^+^ T_EM_ cells. Antigen specific T cell assays and T cell receptor clonality analyses will further educate about the dynamics of SARS-CoV-2 immunological memory formation, which is relevant for the design of future vaccines. Longitudinal investigations within the same patient cohort will be critical to assess the stability of the immune response to SARS-CoV2 infection in correlation with disease severity.

Taken together, this study has highlighted a wide distribution of disease severities in a well characterized and standardized COVID-19 patient cohort despite absence of any relevant previously reported risk factors, in particular old age, comorbidities and therapeutic immunosuppression. Despite the wide spread of disease severity, the immune composition in convalescent patients was fairly stable across disease severity scores except for reduced myeloid cells and naïve T cell frequencies.

## Methods

### Patient characteristics

Patients were randomly selected based on positive SARS-CoV-2-RNA test results by RT-PCR from throat swab samples as well as absence of co-morbidities or pregnancy as assessed by a physician (lung diseases, cardiovascular diseases, cancer, autoimmune diseases, therapeutic immunosuppression). Patient recruitment followed a public announcement for voluntary study participation via the facebook website of the Klinikum r.d. Isar, the University hospital of the Technical University of Munich. Blood sampling occurred 2 weeks after positive SARS-CoV-2-RNA test results in convalescent patients. Detailed patient characteristics are outlined in Figure 1. Clinical COVID-19 scores were assigned to each individual patient according to a questionnaire with a wide range of symptoms (Figure 1) that was established in the presence of a physician. Ethical approval was obtained from the Institutional Review Board of the Technical University of Munich (164/20S and 147/20S). All work involving humans was carried out in accordance with the Declaration of Helsinki for experiments involving humans.

### Cell purification flow cytometry

Peripheral blood mononuclear cells (PBMC) were isolated immediately after blood collection into heparinized collection tubes (within 30 min) by density gradient centrifugation using Biocoll Separting Solution (Merck) and analyzed immediately by flow cytometry (BDFortessa) using the following antibodies (CD45RA-BUV737, clone HI100; CD4-BV785, clone RPA-T4; CD25-BV650, clone BC96; CD8-BV510, clone RPA-T8; CD14-PacificBlue, clone HCD14; CD 19-PerCP-Cy5.5, clone HIB19; CD56-FITC, clone MEM-188; CD197-PE-Cy7, clone G043H7; CD127-PE/Dazzle594, clone A019D5; CD3-PE, clone SK7; γδTCR-APC, clone B1), all from Biolegend. Dead cells were excluded by staining with Hoechst (Sigma-Aldrich).

### Statistical analyses

Error bars indicate the standard error of the mean (SEM); *p* values of 0.05 or less were considered significant. Analyses were performed using GraphPad Prism 8.

## Data Availability

All data of this study is available and archived in the university server.

## Author contributions

CFC, SS, YYC and FS performed the experiments and analyzed the data. C.E.Z. conceived and designed the study and analyzed the data. All authors contributed to writing and reviewing the manuscript.

## Acknowledgements

We thank Andrea Werner and Gustavo Almeida for coordination of the patient recruitment and technical assistance and all members of the Zielinski lab for discussions.

## Notes

### Competing Interest Statement

The authors have declared no competing interest.

### Funding Statement

The authors or their institutions have not at any time received payment or services from a third party for any aspect of the submitted work.

### Author Declarations

Ethical approval was obtained from the Institutional Review Board of the Technical University of Munich (164/20S and 147/20S).

